# Restriction Spectrum Imaging as a quantitative biomarker for prostate cancer with reliable positive predictive value

**DOI:** 10.1101/2024.06.05.24308468

**Authors:** Mariluz Rojo Domingo, Deondre D Do, Christopher C Conlin, Aditya Bagrodia, Tristan Barrett, Madison T Baxter, Matthew Cooperberg, Felix Feng, Michael E Hahn, Mukesh Harisinghani, Gary Hollenberg, Juan Javier-Desloges, Karoline Kallis, Sophia Kamran, Christopher J Kane, Dimitri Kessler, Joshua Kuperman, Kang-Lung Lee, Jonathan Levine, Michael A Liss, Daniel JA Margolis, Ian Matthews, Paul M Murphy, Nabih Nakrour, Michael Ohliger, Courtney Ollison, Thomas Osinski, Anthony James Pamatmat, Isabella R Pompa, Rebecca Rakow-Penner, Jacob L Roberts, Karan Santhosh, Ahmed S Shabaik, Yuze Song, David Song, Clare M. Tempany, Natasha Wehrli, Eric P. Weinberg, Sean Woolen, George Xu, Allison Y Zhong, Anders M Dale, Tyler M Seibert

**Affiliations:** Department of Bioengineering, University of California San Diego, La Jolla, CA, USA; Department of Radiation Medicine, University of California San Diego, La Jolla, CA, USA; Department of Radiology, University of California San Diego, La Jolla, CA, USA; Department of Urology, University of California San Diego, La Jolla, CA, USA; Department of Radiology, University of Cambridge, Cambridge, United Kingdom; Department of Urology, University of California San Francisco, San Francisco, CA, USA; Department of Radiation Oncology, University of California San Francisco, San Francisco, CA, USA; Department of Radiology, Massachusetts General Hospital, Boston, MA, USA; Department of Clinical Imaging Sciences, University of Rochester Medical Center, Rochester, NY, USA; Department of Radiation Oncology, Massachusetts General Hospital, Boston, MA, USA; Department of Urology, University of Texas Health Sciences Center San Antonio, San Antonio, TX, USA; Department of Radiology, Cornell University, Ithaca, NY, USA; Department of Radiology and Biomedical Imaging, University of California San Francisco, San Francisco, CA, USA; Department of Urology, University of Rochester Medical Center, Rochester, NY, USA; Department of Computer Science, University of California San Diego, La Jolla, CA, USA; Department of Pathology, University of California San Diego, La Jolla, CA, USA; Department of Electrical and Computer Engineering, University of California San Diego, La Jolla, CA, USA; Department of Neurosciences, University of California San Diego, La Jolla, CA, USA; Halıcıoğlu Data Science Institute, University of California San Diego, La Jolla, CA, USA

## Abstract

**Background and Objective:** Positive predictive value of PI-RADS for clinically significant prostate cancer (csPCa, grade group [GG]≥2) varies widely between institutions and radiologists. The Restriction Spectrum Imaging restriction score (RSIrs) is a metric derived from diffusion MRI that could be an objectively interpretable biomarker for csPCa.

**Methods:** In patients scanned for suspected or known csPCa at 7 centers, we calculated patient-level csPCa probability based on maximum RSIrs in the prostate, without relying on subjectively defined lesions. We used area under the ROC curve (AUC) to compare patient-level csPCa detection for RSIrs, ADC, and PI-RADS. Finally, we combined RSIrs with clinical risk factors via multivariable regression, training in a single-center cohort and testing in an independent, multi-center dataset.

**Key Findings and Limitations:** Among all patients (n=1892), probability of csPCa increased with higher RSIrs . GG≥4 csPCa was most common in patients with very high RSIrs. Among biopsy-naïve patients (n=877), AUCs for GG≥2 vs. non-csPCa were 0.73 (0.69-0.76), 0.54 (0.50-0.57), and 0.75 (0.71-0.78) for RSIrs, ADC, and PI-RADS, respectively. RSIrs significantly outperformed ADC (*p*<0.01) and was comparable to PI-RADS (*p*=0.31). The combination of RSIrs and PI-RADS outperformed either alone. Combining RSIrs with PI-RADS, age, and PSA density in a multivariable model achieved the best discrimination of csPCa.

**Conclusions and Clinical Implications:** RSIrs is an accurate and reliable quantitative biomarker that performs better than conventional ADC and comparably to expert-defined PI-RADS for patient-level detection of csPCa. RSIrs provides objective estimates of probability of csPCa that do not require radiology expertise.

## Introduction

Multiparametric magnetic resonance imaging (mpMRI) has reduced unnecessary biopsies, decreased overdiagnosis of indolent disease, and improved detection of clinically significant prostate cancer (csPCa, grade group (GG) ≥2)^1–3^. In clinical practice, mpMRI is interpreted qualitatively using the Prostate Imaging Reporting & Data System (PI-RADS v2.1). While negative predictive value (NPV) using PI-RADS is high and fairly consistent^4^, positive predictive value (PPV) for csPCa varies widely across institutions and between radiologists^5,6^. The heavy dependence on user expertise and the variability across readers leads to healthcare disparities by limiting access to high-quality MRI. A quantitative imaging biomarker could help move prostate MRI toward objective interpretation and yield consistent PPV for csPCa-positive biopsy.

Diffusion-weighted MRI is the most important mpMRI sequence for csPCa detection in the PI-RADS system^7^. However, the conventional quantitative metric for diffusion-weighted MRI, apparent diffusion coefficient (ADC), is based on an unrealistically simplistic model that assumes uniform free diffusion of water molecules in the prostate. Restriction Spectrum Imaging (RSI) is a more advanced diffusion-weighted MRI technique that yields a quantitative biomarker (RSI restriction score, or RSIrs) designed to highlight csPCa. In prior retrospective single-center studies, we showed that RSIrs reduced the number of false positives compared to ADC and outperformed ADC for voxel-level and patient-level detection of csPCa^8,9^. A prospective study found that radiation oncologists were much more accurate in outlining csPCa on MRI when using RSIrs than when using conventional MRI alone^10^.

In this study, we evaluate RSIrs as a generalizable tool for patient-level csPCa detection—with objective interpretation—in data from multiple imaging protocols, scanners, vendors, and centers. We also evaluate the accuracy of RSIrs in challenging scenarios: csPCa detection among younger patients^11^ and within the transition zone (TZ)^12,13^. Lastly, we investigate integrating RSIrs with other clinical parameters, such as age and prostate specific antigen density (PSAD) to yield objective estimates of csPCa probability that could serve as a standardized reference for assessing prostate MRIs, independent of radiologist expertise.

## Methods

### Study Population

The data for this study come from seven imaging centers participating in the Quantitative Prostate Imaging Consortium (QPIC): the Center for Translational Imaging and Precision Medicine at the University of California San Diego (CTIPM), UC San Diego Health (UCSD), University of California San Francisco (UCSF), Harvard University affiliated Massachusetts General Hospital (MGH), University of Rochester Medical Center (URMC), University of Texas Health Sciences Center San Antonio (UTHSCSA), and University of Cambridge (Cambridge). The study was approved by each center’s institutional review board (IRB). Data were collected prospectively at UCSD, UTHSCSA, and Cambridge; data were collected retrospectively at the other centers. Participants at UTHSCSA and Cambridge provided written informed consent, while a waiver of consent was approved by the respective IRBs at the other centers for secondary use of routine clinical data. We included individuals aged ≥18 who underwent an MRI for suspected or known csPCa between January of 2016 and March of 2024. Patients were excluded in the event of prior treatment of prostate cancer (PCa) or if there was no available biopsy result from within 6 months of a positive MRI scan (PI-RADS ≥3). Patients with metal implants were also excluded because of the potential to cause significant artifact in MRI. Diagnosis of csPCa was confirmed on biopsy histopathology per clinical routine at each center.

### RSI data acquisition, processing, and modeling

Image post-processing for RSI data included correction for background noise, gradient nonlinearities, and eddy currents^14–16^. Data acquired at CTIPM were also corrected for distortion caused by *B*_0_ inhomogeneity^17^. Automated prostate contours were obtained using an FDA-cleared commercial product (OnQ Prostate, CorTechs.ai, San Diego, CA).

In the RSI framework, diffusion MRI signal is modeled as a combination of exponential decays corresponding to four diffusion microcompartments (intracellular, extracellular, free diffusion, and vascular flow) within each voxel^18^. The RSIrs biomarker is the intracellular signal at a given voxel normalized by median *T*_*2*_-weighted signal in the prostate and multiplied by 1,000 for convenience. RSIrs is highest where intracellular diffusion restriction (hypercellularity) and nucleus-to-cytoplasm ratio are high, both features characteristic of csPCa. Maximum RSIrs is the highest RSIrs value within the prostate^8–10,14,18–20^. Additional details are provided in Supplementary Table 1.

### Patient-level detection of csPCa

For objective and reliable interpretation of MRI results independent of radiologist expertise, risk of csPCa must be determined without subjective lesion delineation. Thus, we assessed csPCa classification performance using maximum RSIrs, which only requires automated segmentation of the prostate. We plotted histograms of maximum RSIrs by csPCa status and obtained the probability (PPV) of csPCa and high-grade csPCa for RSIrs strata by dividing the number of GG≥2 and GG≥3 cases, respectively, by the total number of patients for each bin. Bins spanned 50 RSIrs units, and adjacent bins were combined for illustration purposes if PPV were similar. Pathologic GG is a major prognostic factor for patients with csPCa^21^: GG2 cancer with low-volume Gleason pattern 4 generally poses little risk and may be safely monitored^22^, while GG3-5 cancers are more critical to detect and treat early because of their higher metastatic potential^23^. We showed the GG distribution within each RSIrs stratum among patients who were biopsy-naïve at time of MRI.

To evaluate patient-level detection of csPCa over a range of possible operating points, we plotted the receiver operating characteristic (ROC) curves with csPCa as the outcome of interest. For comparison, we used minimum ADC within the prostate and the highest PI-RADS category for each patient. PI-RADS reporting was performed per clinical routine by experienced, board-certified, fellowship-trained radiologists. We calculated the area under the curve (AUC) with 95% confidence intervals from 10,000-bootstrapping samples and compared bootstrap AUC differences (*α*=0.05). We repeated these analyses stratified by GG (i.e., GG2 vs. non-csPCa, GG3 vs. non-csPCa, etc.). We also evaluated csPCa detection in challenging subsets: patients with TZ lesions and patients with age <60 years.

### Multivariable integrated risk

We used multivariable logistic regression models to combine RSIrs with other routinely available clinical risk factors that physicians may consider in biopsy decisions. We incorporated age, prostate-specific antigen (PSA) level, and PSA density (PSAD). We also evaluated combining these objective variables with expert interpretation of MRI (PI-RADS). Black or African American men are much more likely to develop PCa^24,25^, so we evaluated self-reported race as an additional predictor. We trained the models using UCSD Health data collected on two GE Healthcare Discovery MR750 scanners. The models were tested in remaining patients from all cohorts who were biopsy-naïve at time of MRI and who received a biopsy after MRI. We tested the multivariable models for patient-level detection of csPCa (csPCa vs. non-csPCa) and by GG. We evaluated models with different predictors: (1) age and PSA, which are available before an MRI scan; (2) age and PSAD, which can be computed once MRI is performed; (3) age, PSAD, and RSIrs; (4) RSIrs and PI-RADS, to see if better than either alone; (5) age, PSAD, RSIrs, and PI-RADS; and (6) age, race, PSAD, RSIrs, and PI-RADS.

## Results

### Patient-level detection of csPCa

1892 patients met the criteria for inclusion (Table 1). Data were acquired using 7 distinct acquisition protocols, 2 scanner vendors, 3 scanner models, and 17 MRI scanners (Supplementary Table 2).

**Table 1.** Characteristics of the patients included in this study. *Scans with no PI-RADS available were research-only scans. CTIPM = Center for Translational Imaging and Precision Medicine. UC = University of California. UT = University of Texas. PSA = prostate-specific antigen. PI-RADS = Prostate Imaging Reporting & Data System.

| **Patient Characteristics. Total Study Participants (n = 1892)** |  |
| --- | --- |
| **Cohorts** |  |
| UC San Diego Health | 693 |
| UC San Diego CTIPM | 679 |
| Harvard University's Massachusetts General Hospital (MGH) | 64 |
| University of Rochester Medical Center (URMC) | 251 |
| UC San Francisco (UCSF) | 43 |
| UT Health Sciences Center San Antonio (UTHSCSA) | 147 |
| University of Cambridge | 15 |
| **Clinical Parameters** |  |
| Age (years), median (IQR) | 70 (64-75) |
| PSA (ng/ml), median (SD) | 6.36 (4.65-9.20) |
| Prostate volume (ml), median (IQR) | 51 (36-74) |
| PSA density (ng/ml<sup>2</sup>), median (IQR) | 0.11 (0.07-0.19) |
| **Biopsy** |  |
| Received biopsy prior to MRI scan | 657 |
| Biopsy-naïve at time of MRI scan (had a biopsy within 6 months after MRI) | 1235 (877) |
| **Pathology** |  |
| Systematic biopsy only | 503 |
| Targeted biopsy only | 179 |
| Systematic and targeted biopsy | 710 |
| Prostatectomy | 323 |
| No biopsy within 6 months of MRI scan | 500 |
| **PI-RADS (v2.1)** |  |
| 1 | 636 |
| 2 | 53 |
| 3 | 263 |
| 4 | 453 |
| 5 | 443 |
| Not available* | 44 |
| **Gleason Grade Group** |  |
| Benign | 334 |
| 1 | 296 |
| 2 | 367 |
| 3 | 211 |
| 4 | 81 |
| 5 | 103 |
| **Race & Ethnicity** |  |
| White, Hispanic | 94 |
| White, Non-Hispanic | 1228 |
| White, Ethnicity Other / Unknown | 65 |
| Asian | 120 |
| Black or African American | 117 |

**Table 1.** Characteristics of the patients included in this study.
|  |  |
| --- | --- |
| American Indian/Alaska Native | 6 |
| Native Hawaiian or Other Pacific Islander | 6 |
| Other / Unknown | 256 |

Probability of csPCa increased with higher RSIrs. High-grade (GG4-5) csPCa was proportionally more common among those with highest RSIrs (Figure 1). For RSIrs>500, there was 80% probability of csPCa found on biopsy and 64% probability of GG≥3 PCa, whereas for RSIrs<200, patients had 12% probability of csPCa and only 6% probability of GG≥3 PCa. RSIrs and ADC maps are shown for three representative patients in Figure 2.

**Figure 1.**
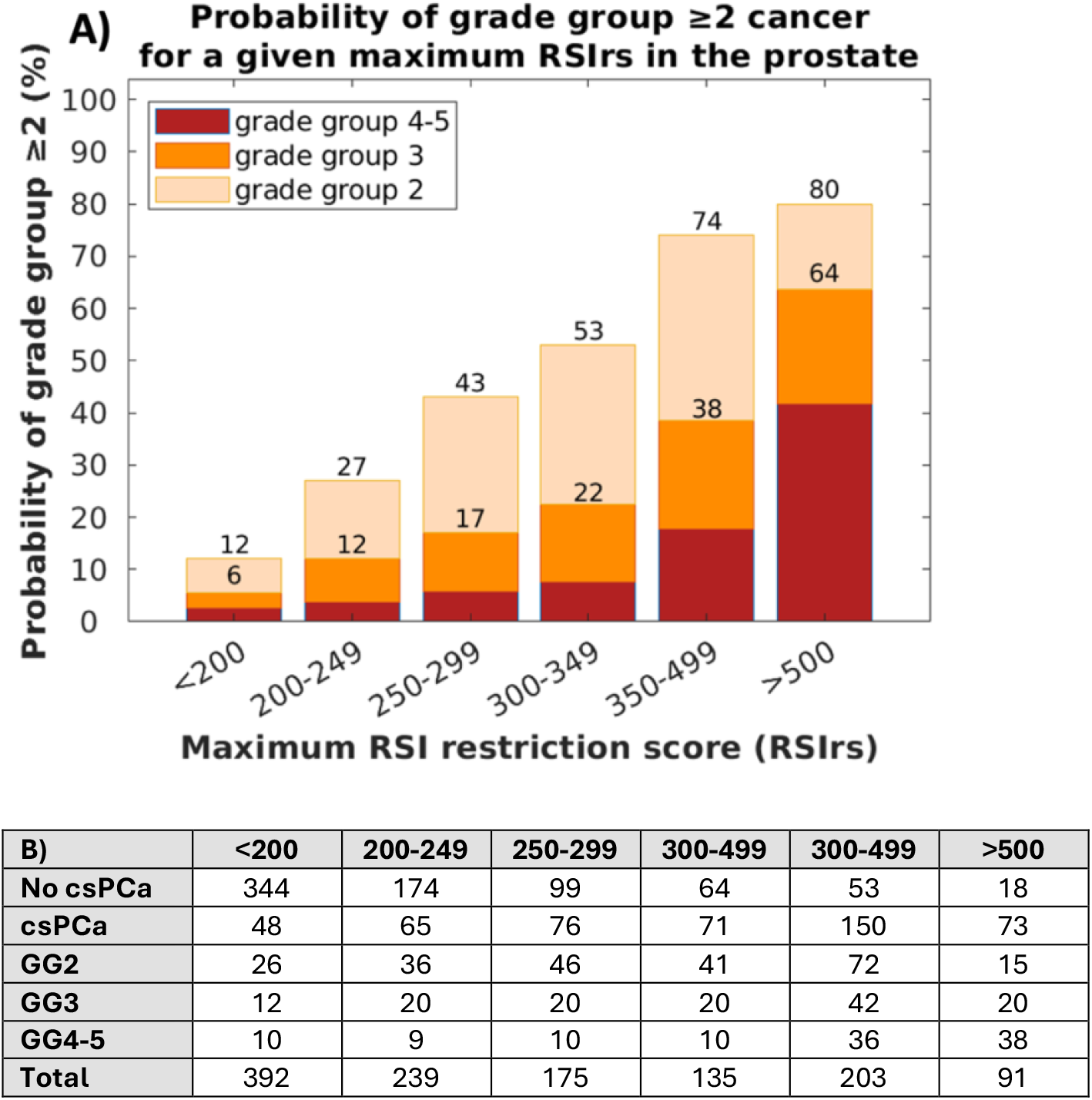
(A) Probability of clinically significant prostate cancer (csPCa) and high-grade csPCa for strata of maximum RSIrs values in data from n=1235 biopsy-naïve patients. The upper number in each column is the probability of csPCa and the lower number is the probability of GG≥3 cancer. (B) Number of patients in each maximum RSIrs stratum in the present dataset. Patients with no biopsy were assumed to not have non-csPCa if expert PI-RADS interpretation was ≤2 and PSA density < 0.15.

**Figure 2.**
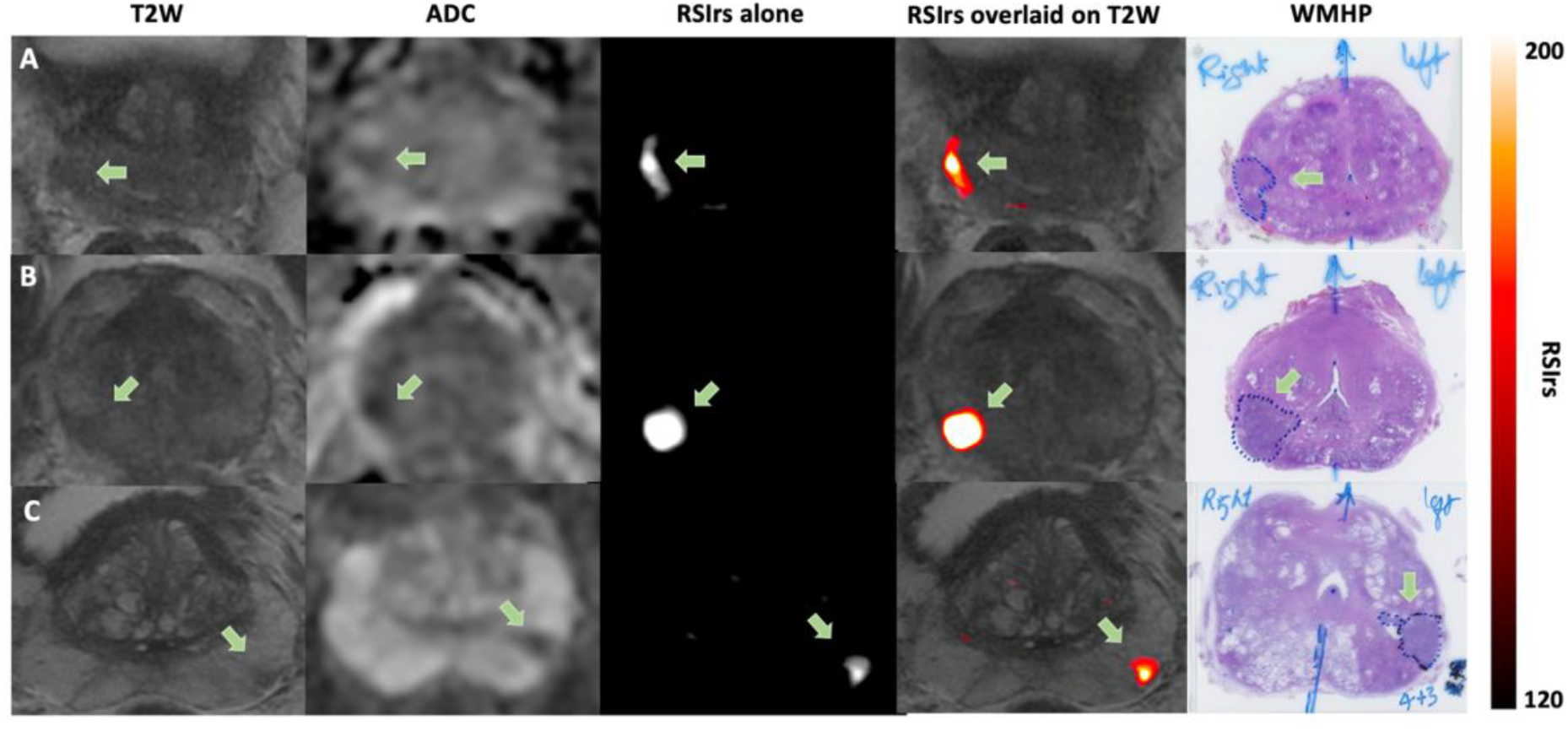
Axial images of *T*_*2*_-weighted (T2W) MRI, conventional ADC, RSIrs, RSIrs overlaid on the anatomical T2W images, and whole-mount histopathology for three representative patients who underwent radical prostatectomy within 6 months of MRI. The RSI maps highlight the areas where clinically significant prostate cancer (csPCa) was confirmed on whole-mount histopathology. All three patients had PI-RADS 4 lesions in the peripheral zone (green arrows). Prostatectomy results showed that patient A had Gleason 4+4 prostate cancer (grade group 4), while patients B and C had Gleason 4+3 cancer (grade group 3).

ROC curve analysis demonstrated that RSIrs was superior to ADC and comparable to PI-RADS for patient-level detection of csPCa (Figure 3). Among 877 biopsy-naïve patients who underwent biopsy after MRI, median AUC for GG≥2 vs. non-csPCa was 0.73 (0.69-0.76) for RSIrs, 0.54 (0.50-0.57) for ADC, and 0.75 (0.71-0.78) for PI-RADS. RSIrs significantly outperformed ADC (*p*<0.01) and was comparable to PI-RADS (*p*=0.31).

**Figure 3.**
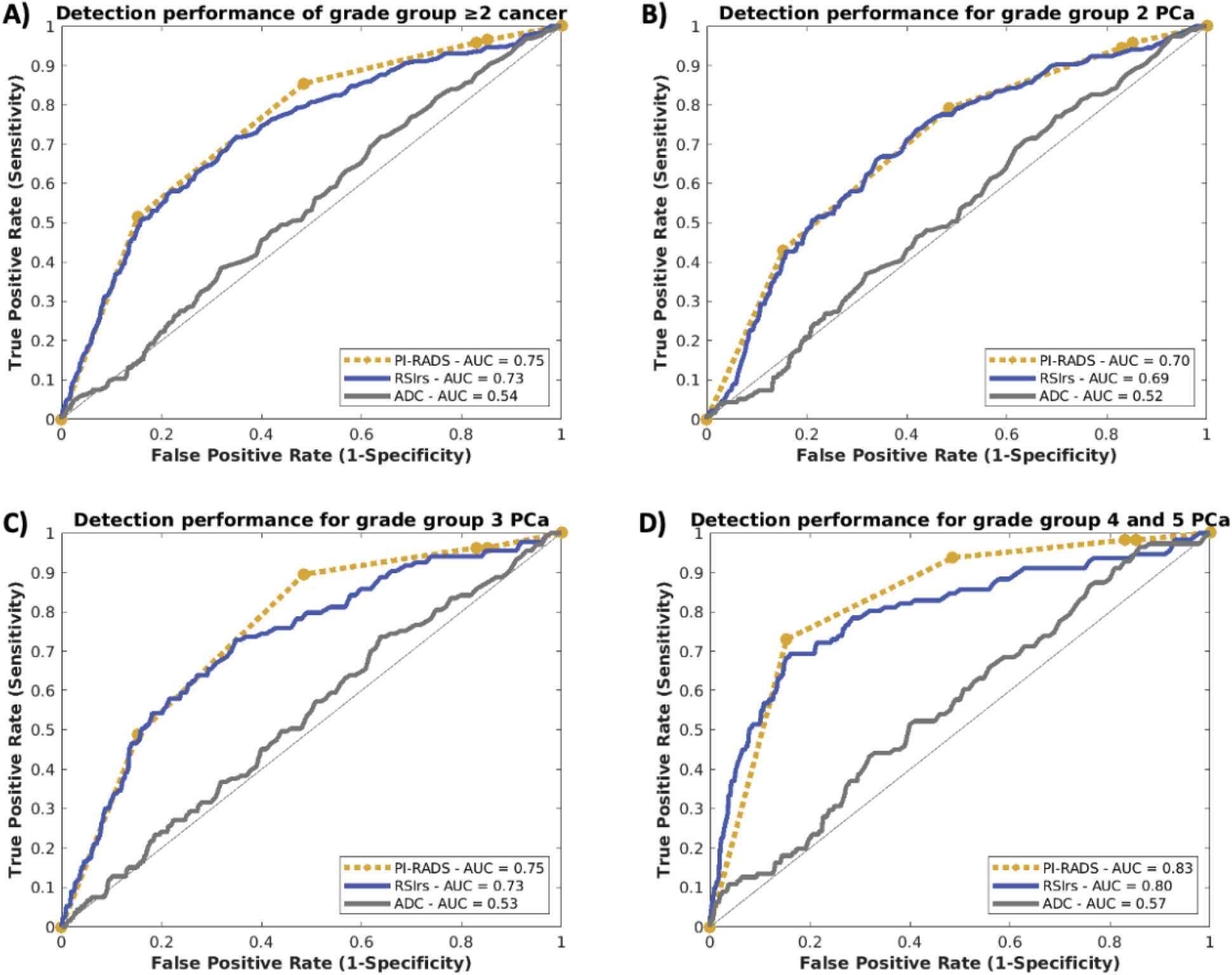
ROC curves by grade group (GG) for patient-level detection of csPCa using RSIrs, PI-RADS and ADC. Patients were included if they were biopsy-naïve at time of MRI and underwent biopsy after MRI. Yellow circles correspond to PI-RADS thresholds. A) AUCs for discrimination of GG ≥2 PCa vs no csPCa (n=877). B) AUCs for GG2 PCa detection vs no csPCa (n=633). C) AUCs for GG3 PCa detection vs no csPCa (n=531). D) AUCs for GG4-5 PCa detection vs no csPCa (n=509). RSIrs was superior to ADC in detection of GG≥2 PCa, GG2, GG3 and GG4-5 (*p*<0.01). AUCs with 95% confidence intervals and *p*-values are in Supplementary Table 3.

When comparing GG≥3 to non-csPCa (i.e., excluding GG2), median AUCs were 0.76 (0.72-0.80) for RSIrs, 0.55 (0.50-0.60) for ADC, and 0.79 (0.76-0.82) for PI-RADS. RSIrs significantly outperformed ADC (*p*<0.01) and was comparable to PI-RADS (*p*=0.14). Both RSIrs and PI-RADS showed partial specificity for high-grade csPCa, with higher performance for detection of GG3 and GG4-5 than for GG2 (Figure 3 and Supplementary Table 3).

RSIrs performed similarly to expert PI-RADS in patients with lesions in the TZ (*p*=0.90) and PZ (*p*=0.07). RSIrs performed similarly to PI-RADS in patients <60 years (*p*=0.12) and >60 years (*p*=0.11). Subset analyses by race were limited by small sample size for most groups (Supplementary Table 3).

### Multivariable integrated risk

Models to combine predictor variables were trained in 554 patients, including 232 with no biopsy but presumed free of csPCa (PI-RADS 1-2 and PSAD≤0.15)^26^ and excluding patients that did not have a PI-RADS score available. Models were tested in an independent dataset from multiple institutions with 664 patients, all biopsy naïve before MRI and with biopsy confirmation of csPCa status. The combination of RSIrs and PI-RADS outperformed either alone (*p*<0.01 and *p*=0.01, respectively), and a model of age, PSAD, PI-RADS, and RSIrs achieved the best discrimination of csPCa, outperforming RSIrs alone and PI-RADS alone (*p*<0.01; Table 2). Addition of race did not significantly improve performance in any of the multivariable models.

**Table 2.** Results from the multivariable logistic regression models for combinations of RSIrs with clinical and imaging parameters for discrimination of clinically significant prostate cancer (csPCa, grade group [GG] ≥ 2). Group A) independent testing in all biopsy-naïve patients at time of MRI with biopsy confirmed diagnosis who were not used for training (n=664); comparison is csPCa vs. no csPCa (benign or grade group 1, GG1). Group B) GG2 vs. non-csPCa: subset of independent testing dataset with either GG2 csPCa or no csPCa (n=500). Group C) GG3 vs. non-csPCa: subset of independent testing dataset with either GG3 csPCa or no csPCa (n=409). Group D) GG4-5 vs. no csPCa: subset of independent testing dataset with GG4 csPCa, GG5 csPCa, or no csPCa (n=393). 95% confidence intervals were calculated from 10,000-bootsrapping stratified by csPCa.

|  | **AUCs for csPCa discrimination in multivariable models** |  |  |  |
| --- | --- | --- | --- | --- |
| **Test Group** | **RSIrs** | **PI-RADS** | **ADC** | **RSIrs, PI-RADS\*** |
| **A) GG ≥ 2 cancer** | 0.72 [0.68-0.76] | 0.74 [0.70-0.77] | 0.54 [0.50-0.59] | 0.77 [0.73-0.81] |
| **B) GG2 cancer** | 0.69 [0.64-0.74] | 0.69 [0.64-0.73] | 0.53 [0.50-0.58] | 0.73 [0.68-0.77] |
| **C) GG3 cancer** | 0.72 [0.66-0.78] | 0.74 [0.69-0.79] | 0.54 [0.50-0.60] | 0.78 [0.72-0.83] |
| **D) GG4-5 cancer** | 0.80 [0.73-0.86] | 0.85 [0.80-0.89] | 0.58 [0.51-0.65] | 0.87 [0.82-0.92] |
|  | **PSA, Age** | **PSAD, Age, RSIrs** | **PSAD, Age, RSIrs, PI-RADS\*** | **PSAD, Age, Race, RSIrs, PI-RADS\*** |
| **A) GG ≥ 2 cancer** | 0.61 [0.56-0.66] | 0.74 [0.70-0.77] | 0.78 [0.74-0.82] | 0.78 [0.74-0.82] |
| **B) GG2 cancer** | 0.53 [0.50-0.58] | 0.69 [0.64-0.74] | 0.73 [0.69-0.78] | 0.74 [0.69-0.78] |
| **C) GG3 cancer** | 0.65 [0.57-0.72] | 0.73 [0.67-0.79] | 0.78 [0.72-0.83] | 0.78 [0.72-0.83] |
| **D) GG4-5 cancer** | 0.77 [0.70-0.83] | 0.86 [0.81-0.91] | 0.90 [0.85-0.94] | 0.90 [0.85-0.94] |
| PSAD: prostate specific antigen density |  |  |  |  |
| *: Performance in predictor groups marked with an asterisk is significantly better than that of RSIrs. |  |  |  |  |

## Discussion

We assessed RSIrs as an objective MRI biomarker for detecting csPCa at the patient level. In contrast to ADC, which typically becomes clinically useful after a radiologist identifies a suspicious lesion, RSIrs assessed automatically within the entire prostate performed comparably to expert PI-RADS for patient-level detection of csPCa in a large, heterogenous and multi-center dataset. Moreover, RSIrs performs best for the high-grade cancers that are also most important to detect. An automated measurement of RSIrs can give physicians and patients an objective and reliable estimate of the likelihood of csPCa or high-grade csPCa.

Subspecialist radiologists are often at elite centers that provide care for only a small proportion of patients. A quantitative biomarker could contribute to making accurate prostate MRI accessible to patients who do not receive their care at these elite centers. The PPV of RSIrs is inherently reproducible for a given scan, as it is calculated objectively from the MRI. Use of RSIrs, then, could make prostate MRI more reliable and more readily interpretable for referring physicians and their patients. By addressing the variable PPV of PI-RADS and reducing dependence on reader expertise, implementation of objective biomarkers could increase health equity in the PCa diagnostic pathway.

RSIrs requires only an RSI MRI acquisition lasting 2-3 minutes and a *T*_*2*_-weighted MRI acquisition (another 2-3 minutes). Thus, 4-6 minutes of scan time can yield an automated RSIrs biomarker with performance comparable to expert radiologists’ evaluations of a full PI-RADS mpMRI scan. RSI acquisitions are compatible with standard clinical scanners and do not require administration of intravenous contrast. Installing the RSI acquisition protocols on modern scanners involves simply saving protocol files on the scanner. Calculation of RSIrs can be performed by software on a desktop personal computer.

Maximum RSIrs is a quantitative biomarker that is readily interpretable. Thus, use of RSIrs could establish a floor for performance of csPCa detection regardless of available radiology expertise. Radiologist interpretation of MRI remains important for evaluation of secondary questions: extraprostatic extension, tumor proximity to the neurovascular bundles, and seminal vesicle involvement. However, these latter questions are mostly relevant only after a biopsy-confirmed diagnosis of csPCa is established and therefore apply to a smaller subset of patients. For the initial question of whether csPCa is likely to be found on biopsy, RSIrs performs comparably to expert-defined PI-RADS, and the combination of both is better than either alone.

We focused this study on patient-level csPCa detection. Another important role of MRI is tumor localization for targeted biopsy^27–32^and radiotherapy planning^33–35^. Radiologist-defined lesion segmentations were not available to perform lesion-level analysis of this large dataset. The commercial software our centers use to delineate biopsy targets does not permit export of those segmentations and automatically deletes them to make room for future studies. In any event, expert-defined lesions are subjectively identified, thus undermining the primary goal of the study to consider approaches independent of radiologist expertise. Prior work, though, has shown that RSIrs maps are useful for localization of csPCa (Figure 2). There is a strong correlation between RSI and csPCa on whole-mount histopathology, and RSIrs is superior to ADC for voxel-level detection of csPCa^9,36^. Further, a prospective study evaluated radiation oncologists’ ability to delineate csPCa on MRI; these non-radiologists were much more accurate when using RSIrs maps vs. conventional MRI alone, confirming that RSIrs maps reflect the location of csPCa and make it more apparent to non-experts^10^.

Automated and quantitative MRI approaches may help alleviate the growing shortage of expert radiologists relative to an anticipated surge in PCa diagnoses^37^. Other MRI biomarkers have also shown potential clinical utility in prior studies^38,39^. To our knowledge, the present study is the largest and most comprehensive validation of a quantitative MRI biomarker for patient-level csPCa detection. Ongoing research evaluates whether incorporating RSIrs into radiomics-based analysis and deep-learning artificial intelligence tools could further enhance detection performance.

Our study has some limitations. First, biopsy techniques are prone to sampling error and therefore represent an imperfect gold standard. Nonetheless, most patients here underwent both systematic and targeted biopsy, which is the current clinical standard and captures most csPCa^4,28,29,40^. Consistent with clinical guidelines, patients with non-suspicious prostate MRI typically did not undergo biopsy, raising the possibility of false negatives on PI-RADS, though the risk of this is low^4^. Also, patients with hip implants were excluded from this study; the effect of metal artifact on RSIrs is the subject of ongoing research.

## Conclusions

In heterogeneous data from multiple imaging centers, RSIrs proved to be a quantitative imaging biomarker that performs comparably to expert-defined PI-RADS for patient-level detection of csPCa. With only 4-6 minutes of scan time on standard clinical MRI platforms, RSIrs gives objective estimates of probability of csPCa, thus addressing the current clinical challenge of unreliable PPV with PI-RADS.

## Data Availability

All data produced in the present study are available upon reasonable request to the authors

## Supplementary Material

**Supplementary Figure 1.**
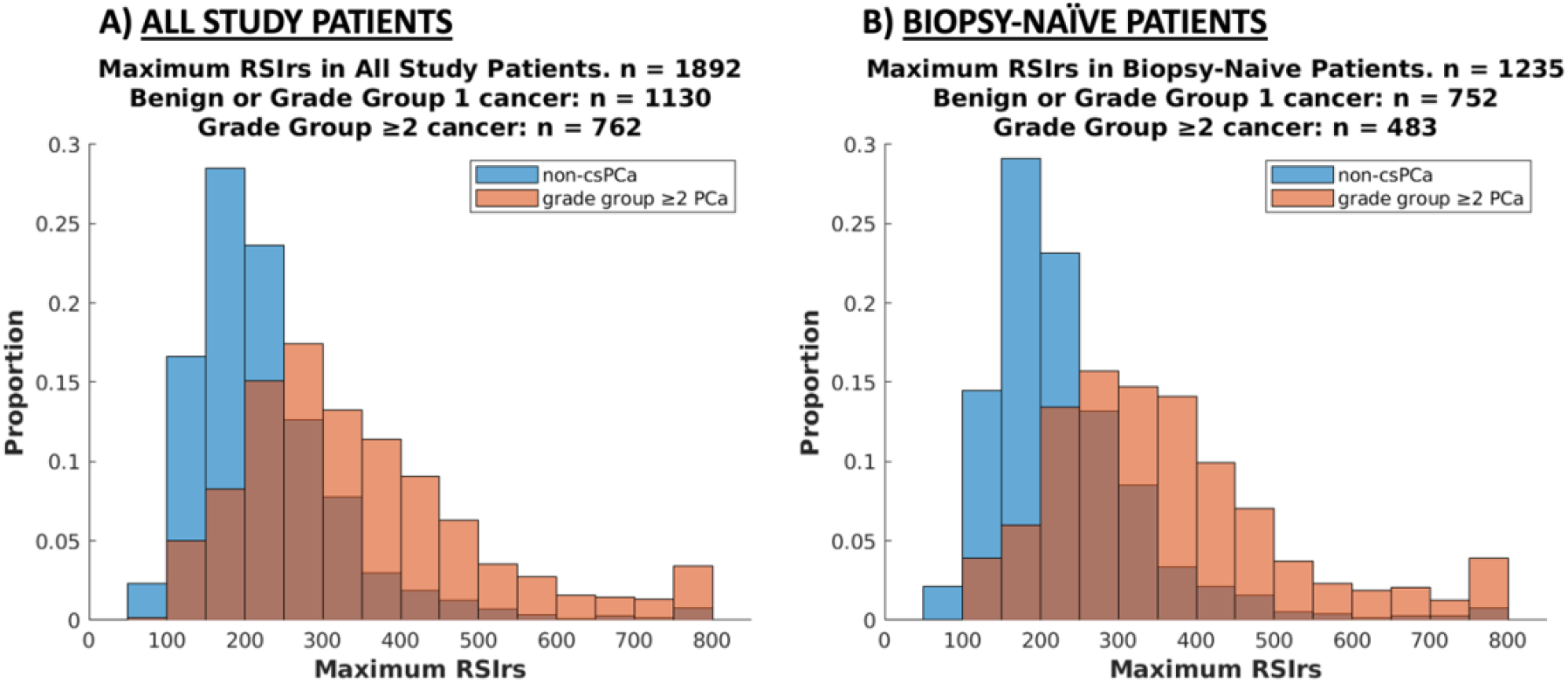
A) Histogram of RSIrs values in all patients in the study. B) Histogram of RSIrs values in patients who were biopsy-naïve at time of MRI.

**Supplementary Table 1.** The RSI model computes the sum of DWI signal from the four compartments as expressed by the formula above. *S(b)* represents the measured diffusion-weighted imaging (DWI) signal intensity at a specific *b*-value. The signal is modeled as a linear combination of exponential decays, each corresponding to one of four diffusion compartments. *C*_*i*_ denotes the signal contribution of a particular compartment to the overall signal; these contributions are determined through model-fitting. The diffusion coefficients, *D*_*i*_ , are set to empirically determined values for each of the four tissue compartments (*C*_*i*_ ).

| RSI model formula | RSI compartment | Fixed Diffusion Coefficient (s/mm<sup>2</sup>) |
| --- | --- | --- |
| $S(b) = \sum_{i=1}^4 C_i e^{-bD_i}$ | Restricted Diffusion ($C_1$) | $1.1 * 10^{-4} (D_1)$ |
| | Hindered Diffusion ($C_2$) | $1.8 * 10^{-3} (D_2)$ |
| | Free Diffusion ($C_3$) | $3.6 * 10^{-3} (D_3)$ |
| | Vascular Flow ($C_4$) | $0.1220 (D_4)$ |

**Supplementary Table 2.** MRI acquisition parameters for each cohort. Parameters such as echo time (TE), repetition time (TR), matrix acquisition or slice thickness differ between RSI protocols. FOV: field-of-view. FSE: fast spin echo. EPI: echo-planar imaging. DWI: diffusion-weighted imaging. UCSD: University of California San Diego. CTIPM: Center for Translational Imaging and Precision Medicine. MGH: Harvard University’s Massachusetts General Hospital. URMC: University of Rochester Medical Center. UTHSCSA: University of Texas Health Sciences Center San Antonio. UCSF: University of California San Francisco. Cambridge: University of Cambridge.

| UCSD CTIPM | DWI | $T_2$-weighted |
| --- | --- | --- |
| Pulse sequence | Diffusion-weighted EPI | Fast Spin Echo (FSE) |
| TR (ms) | 4500 | 7000 |
| TE (ms) | 69 | 100 |
| FOV (mm) | 240 x 120 | 240 x 240 |
| Matrix<br>[resampled dimensions] | 96 x 48 [128 x 128] | 320 x 320 [512 x 512] |
| Slices | 16 | 32 |
| Slice Thickness (mm) | 6 | 3 |
| $b$-values (s/mm<sup>2</sup>)<br>[number of samples] | 0[1], 500[6],<br>1000[6], 2000[12] | N/A |
| Field Strength (T) | 3 | 3 |

| UCSD Health | DWI | $T_2$-weighted |
| --- | --- | --- |
| Pulse sequence | Diffusion-weighted EPI | Fast Spin Echo (FSE) |
| TR (ms) | 4000 | 5300 |
| TE (ms) | 69 | 100 |
| FOV (mm) | 240 x 120 | 200 x 200 |
| Matrix<br>[resampled dimensions] | 96 x 48 [256 x 256] | 320 x 320 [512 x 512] |
| Slices | 16 | 32 |
| Slice Thickness (mm) | 6 | 3 |
| $b$-values (s/mm<sup>2</sup>)<br>[number of samples] | 0[1], 500[8],<br>1000[8], 2000[16] | N/A |
| Field Strength (T) | 3 | 3 |

| URMC | DWI | $T_2$-weighted |
| --- | --- | --- |
| Pulse sequence | Diffusion-weighted EPI | Fast Spin Echo (FSE) |
| TR (ms) | 3800 | 4800 |
| TE (ms) | 85 | 104 |
| FOV (mm) | 52 x 52 | 180x180 |
| Matrix<br>[resampled dimensions] | 100 x 52 [104 x 200] | 384 x 365 [384 x 384] |
| Slices | 22 | 32 |
| Slice Thickness (mm) | 4 | 3 |
| $b$-values (s/mm<sup>2</sup>)<br>[number of samples] | 0[1], 500[6],<br>1000[6], 2000[6] | N/A |
| Field Strength (T) | 3 | 3 |

| MGH | DWI | $T_2$-weighted |
| --- | --- | --- |
| Pulse sequence | Diffusion-weighted EPI | Fast Spin Echo (FSE) |
| TR (ms) | 4500 | 3937 |
| TE (ms) | 59 | 169 |
| FOV (mm) | 240 x 120 | 160 x 160 |
| Matrix<br>[resampled dimensions] | 96 x 48 [128x128] | 360 x 224<br>[1024 x 1024] |
| Slices | 16 | 40 |
| Slice Thickness (mm) | 6 | 3 |
| $b$-values (s/mm<sup>2</sup>)<br>[number of samples] | 0[1], 500[6],<br>1000[6], 2000[12] | N/A |
| Field Strength (T) | 3 | 3 |

| Cambridge | DWI | $T_2$-weighted |
| --- | --- | --- |
| Pulse sequence | Diffusion-weighted EPI | Fast Spin Echo (FSE) |
| TR (ms) | 4500 | 3130 |
| TE (ms) | 68 | 98 |
| FOV (mm) | 220 x 110 | 180 x 180 |
| Matrix<br>[resampled dimensions] | 96 x 48 [256 x 256] | 448 x 256 [512 x 512] |
| Slices | 8 | 30 |
| Slice Thickness (mm) | 4 | 3 |
| *b*-values (s/mm<sup>2</sup>)<br>[number of samples] | 0[1], 500[2],<br>1000[2], 2000[4] | N/A |
| Field Strength (T) | 3 | 3 |

| UTHSCSA | DWI | *T*<sub>2</sub>-weighted |
| --- | --- | --- |
| Pulse sequence | Diffusion-weighted EPI | Fast Spin Echo (FSE) |
| TR (ms) | 6300 | 4710 |
| TE (ms) | 105 | 100 |
| FOV (mm) | 52 x 100 | 180 x 180 |
| Matrix<br>[resampled dimensions] | 52 x 100 x [104 x 200] | 240 x 320 x [320 x 320] |
| Slices | 22 | 30 |
| Slice Thickness (mm) | 4 | 3 |
| *b*-values (s/mm<sup>2</sup>)<br>[number of samples] | 0[1], 500[6],<br>1000[18], 2000[42] | N/A |
| Field Strength (T) | 3 | 3 |

| UCSF | DWI | *T*<sub>2</sub>-weighted |
| --- | --- | --- |
| Pulse sequence | Diffusion-weighted EPI | Fast Spin Echo (FSE) |
| TR (ms) | 4500 | 2964 |
| TE (ms) | 73 | 150 |
| FOV (mm) | 200 x 200 | 220 x 220 |
| Matrix<br>[resampled dimensions] | 256 x 256 [256 x 256] | 512 x 512 [320 x 320] |
| Slices | 35 | 36 |
| Slice Thickness (mm) | 3 | 3 |
| *b*-values (s/mm<sup>2</sup>)<br>[number of samples] | 0[5], 100[6], 800[12],<br>1400[12], 2500[18] | N/A |
| Field Strength (T) | 3 | 3 |

**Supplementary Table 2.** MRI acquisition parameters for each cohort.
| Institution | Scanner models | Number of Stations |
| --- | --- | --- |
| UCSD CTIPM | GE Healthcare Discovery MR750,<br>GE Healthcare Signa Premier | 4 |
| UCSD Health | GE Healthcare Discovery MR750,<br>GE Healthcare Signa Premier | 4 |
| URMC | SIEMENS Skyra | 2 |
| MGH | GE Healthcare Signa Premier | 1 |
| UCSF | GE Healthcare Signa Premier | 2 |
| Cambridge | GE Healthcare Discovery MR750 | 1 |
| UTHSCSA | SIEMENS Skyra | 3 |
| Total | 3 scanner models | 17 stations |

**Supplementary Table 3.** AUC values for RSIrs, PI-RADS and ADC in different subsets. 95% confidence intervals were calculated from 10,000-bootsrapping stratified by grade group. All cohorts are against non-csPCa (benign and grade group 1). GG: Grade Group.

| Patient Cohort | n | % with csPCa | AUC RSIRs | AUC PI-RADS | AUC ADC |
| --- | --- | --- | --- | --- | --- |
| GG $\geq 2$ | 877 | 55 | 0.73 [0.69,0.76] | 0.75 [0.71,0.78] | 0.54 [0.50,0.57] |
| GG $\geq 3$ | 642 | 38 | 0.76 [0.72,0.80] | 0.79 [0.76,0.82] | 0.55 [0.50,0.60] |
| GG 2 only | 633 | 37 | 0.69 [0.65,0.73] | 0.70 [0.66,0.74] | 0.52 [0.48,0.57] |
| GG 3 only | 531 | 25 | 0.73 [0.68,0.77] | 0.75 [0.71,0.80] | 0.53 [0.48,0.59] |
| GG 4-5 only | 509 | 22 | 0.80 [0.74,0.85] | 0.83 [0.79,0.87] | 0.57 [0.51,0.63] |
| TZ csPCa | 329 | 43 | 0.72 [0.66,0.78] | 0.72 [0.66,0.77] | 0.57 [0.51,0.63] |
| PZ csPCa | 602 | 55 | 0.72 [0.68,0.76] | 0.76 [0.73,0.80] | 0.52 [0.48,0.57] |
| < 60 years old | 103 | 45 | 0.79 [0.69,0.87] | 0.70 [0.60,0.79] | 0.57 [0.45,0.68] |
| $\geq 60$ years old | 772 | 56 | 0.72 [0.68,0.75] | 0.75 [0.72,0.78] | 0.53 [0.49,0.57] |
| Race White | 672 | 53 | 0.74 [0.70,0.77] | 0.76 [0.72,0.79] | 0.54 [0.49,0.58] |
| Race Asian | 47 | 64 | 0.74 [0.59,0.87] | 0.78 [0.64,0.89] | 0.62 [0.44,0.79] |
| Race Black | 51 | 45 | 0.68 [0.53,0.82] | 0.60 [0.46,0.74] | 0.54 [0.38,0.70] |
| Race Other / Unknown | 107 | 64 | 0.70 [0.59,0.80] | 0.72 [0.62,0.80] | 0.57 [0.45,0.68] |

**Supplementary Table 3.** AUC values for RSIRs, PI-RADS and ADC in different subsets. 95% confidence intervals were calculated from 10,000-bootstraping stratified by grade group. All cohorts are against non-csPCa (benign and grade group 1). GG: Grade Group.
|  | *p*-value RSIRs vs. PI-RADS | *p*-value RSIRs vs. ADC |
| --- | --- | --- |
| GG $\geq 2$ | 0.31 | <0.01 |
| GG $\geq 3$ | 0.14 | <0.01 |
| GG 2 only | 0.75 | <0.01 |
| GG 3 only | 0.31 | <0.01 |
| GG 4-5 only | 0.20 | <0.01 |
| TZ csPCa | 0.90 | <0.01 |
| PZ csPCa | 0.07 | <0.01 |
| < 60 years old | 0.12 | <0.01 |
| $\geq 60$ years old | 0.11 | <0.01 |
| Race White | 0.32 | <0.01 |
| Race Asian | 0.61 | 0.28 |
| Race Black | 0.41 | 0.12 |
| Race Other / Unknown | 0.82 | 0.05 |

